# Evaluating Clinical Presentation and Long-Term Outcomes in Individuals with Genetic and Non-Genetic Epilepsy Treated with Epilepsy Surgery: A Single-Center Study

**DOI:** 10.1101/2023.12.03.23299306

**Authors:** Alina Ivaniuk, Christian M Boßelmann, Elia Pestana-Knight, Xiaoming Zhang, Lara Jehi, William Bingaman, Imad Najm, Dennis Lal

## Abstract

**Background:** Many genetic epilepsy disorders are characterized by focal epilepsy or exhibit focal or lateralized features, which make individuals with such conditions potential surgical candidates. However, surgical outcomes in epilepsy caused by germline genetic variants and value of genetic testing in presurgical assessment remains unclear.

**Methods:** This retrospective cross-sectional study included people with germline genetic epilepsy and non-genetic epilepsy identified among 2879 people with epilepsy who underwent resective surgery or laser ablation at Cleveland Clinic in years 1997-2022. We separated individuals with germline genetic epilepsies into mTORopathies (clinically or genetically diagnosed tuberous sclerosis and genetically diagnosed GATOR-related epilepsies) and other genetic etiologies. We compared clinical variables between the groups using Kruskal-Wallis and Fisher tests as appropriate. We analyzed cumulative seizure recurrence risk over 5 years with Kaplan-Meier curves and used Cox proportional hazards model for univariate and multivariate analysis. We repeated the analysis after matching both genetic sub-groups to non-genetic group 1:1 by propensity scores estimated by a generalized linear model.

**Results:** We included 49 individuals with genetic epilepsies (32 individuals with mTORopathies and 17 individuals with other genetic etiologies) and 585 individuals with non-genetic epilepsy. Individuals with genetic epilepsies in both groups were younger at seizure onset (p<0.001) and at surgery (p<0.001), had a higher preoperative seizure frequency (p<0.001), higher rate of developmental delay (p<0.001), lower rate of focal interictal (p=0.005) and ictal (p<0.001) findings on EEG, and more extratemporal and extensive resections (p<0.001). At the last follow-up, individuals with other genetic etiologies had a lower rate of seizure freedom than individuals with mTORopathies and non-genetic etiologies; still, 70.6% achieved seizure reduction. Individuals with other genetic etiologies had a higher cumulative risk of seizure recurrence at 5 years both in univariate (HR 3.07, 95% CI 1.4-6.74) and multivariate (HR 3.81, 95% CI 1.69-8.62, p=0.001) analyses, which was reproducible in a matched cohort analysis.

**Conclusions:** People with germline genetic epilepsy across multiple etiologies could be surgical candidates. However, seizure recurrence may be faster in genetic epilepsies other than mTORopathies. Further studies are required to clarify surgical candidacy in different genetic epilepsy disorders and establish value of presurgical genetic testing.

## Introduction

In one-third of individuals with focal epilepsy, seizures are not controlled with anti-seizure medications despite the availability of newer agents.^1^ For appropriately selected medication-resistant individuals, epilepsy surgery presents a safe and effective treatment option. Well-defined epileptogenic zone and the presence of a corresponding lesion on MRI have been associated with good surgical outcomes across different studies and are thus major determinants of surgical candidacy.^2,3^

With the wide adoption of clinical genetic testing, an underlying genetic cause could be identified for a range of seizure disorders.^4,5^ In some of them, focal epilepsy is a main phenotypic feature. Examples of such disorders include mTORopathies – conditions caused by pathogenic variants affecting genes that encode regulators of mechanistic target of rapamycin complex 1. Most common mTORopathies include tuberous sclerosis caused by germline pathogenic variants in *TSC1* and *TSC2* genes and focal lesional and non-lesional epilepsies caused by germline pathogenic variants in *DEDPC5* and *NPRL2*/*NPRL3* genes encoding subunits of the GATOR1 complex.^6^ In both tuberous sclerosis and GATOR1-related epilepsies, epilepsy and neuropsychiatric symptoms are attributed to malformations of cortical development – tubers and focal cortical dysplasias, respectively – which share common neuropathological findings of cytomegaly, abnormal cortical lamination, and hyperexcitability.^7–10^ Tuberous sclerosis can have a favorable prognosis depending on the lesion burden, completeness of resection of epileptogenic malformation, and focality of preoperative EEG findings.^7,11–13^ Likewise, existing evidence on GATOR1-related disorders indicates good postsurgical seizure outcomes in cases with well-defined focal lesions where complete resection is possible.^8,14–16^

Genetic epilepsies and epilepsy syndromes traditionally expected to have multifocal or generalized seizures, such as channelopathies or synaptopathies, may exhibit focal or lateralizing features, and thus be considered surgical candidates in specific clinical scenarios.^17,18^ However, evidence on success of epilepsy surgery in such cases is limited and heterogeneous. For example, epilepsy in Dravet syndrome, a developmental and epileptic encephalopathy caused by loss-of-function pathogenic variants in *SCN1A*, is characterized by generalized and hemiclonic seizures at the onset, with focal seizures possible later in the course of the disease.^19–21^ Another *SCN1A*-related seizure phenotype, early-onset developmental and epileptic encephalopathy caused by gain-of-function variants, presents with focal seizures.^22^ Several available reports of epilepsy surgery in *SCN1A*-related epilepsy suggest discouraging results.^23,24^ Malformations of cortical development are reported in *PCDH19*-related epilepsy, in which the core phenotype constitutes generalized and focal seizures, intellectual disability, and autism spectrum disorder; several case series with favorable outcomes in this disorder have been reported.^25,26^

Therefore, people with genetic epilepsy could be considered surgical candidates in specific clinical scenarios. Despite becoming an increasingly common clinical variable in presurgical assessment of individuals with epilepsy, data on the role of genetic etiology in the evaluation for epilepsy surgery and its impact on postsurgical outcomes is still limited to descriptive evidence drawn from case series and small cohort reports.^24,27,28^ No studies have compared seizure outcomes in individuals with genetic epilepsy and non-genetic epilepsy. Since individuals with genetic epilepsy considered to be surgical candidates are assessed presurgically with the same tools and methods as individuals with non-genetic epilepsy, comparing data on postsurgical outcomes between these two groups appears logical, would facilitate surgical decision-making, and could provide background for the development of clinical recommendations.

Here, we conduct a comparative assessment of the clinical data and outcomes of 49 individuals with mTORopathies and other genetic etiologies who underwent epilepsy surgery at the Cleveland Clinic Epilepsy Center with individuals with non-genetic causes of epilepsy. Since the differences in clinical presentation of individuals with genetic and non-genetic epilepsies are expected and could affect the outcomes, we used multivariate analysis and propensity score matching to mitigate the bias and show reproducibility of results with two different statistical approaches. Our study improves preliminary evidence on the feasibility of surgical treatment in individuals with genetic epilepsy and highlights clinical features that may prompt consideration for genetic testing in presurgical evaluation.

## Methods

### Cohort acquisition

This retrospective cross-sectional cohort study included 2879 people with epilepsy who received epilepsy surgery at the Cleveland Clinic Health System of the Cleveland Clinic Foundation (CCF) from 1997 to 2022. We included individuals who underwent resective epilepsy surgery or lesion ablation. In case an individual received several surgeries, we only considered the outcome and EEG-related variables pertaining to the first surgery. We excluded individuals who i) Underwent neurostimulation or callosotomy only and ii) Had less than one year of follow-up as of December 31, 2022.

We identified individuals with potentially germline genetic epilepsy by querying electronic medical records (EMR) for entries containing names of genes associated with epilepsy (defined by Macnee et al.^5^) and/or coded with International Classification of Diseases – 10^th^ Revision codes Q90-99 (“Chromosomal abnormalities, not elsewhere classified”). We further refer to epilepsy caused by germline genetic variants as genetic epilepsy. We matched retrieved patient identifiers with the operating room records of the Epilepsy Center to identify a group of patients who underwent epilepsy surgery. After retrieving individuals with potentially genetic epilepsy treated with epilepsy surgery, we reviewed their medical records to verify the genetic diagnosis. Individuals with tuberous sclerosis complex (TSC) were included if they met definite TSC criteria, either genetic or clinical.^29^ For genetic diagnoses other than TSC, we required the presence of a pathogenic/likely pathogenic variant on clinical genetic testing. All genetic findings were reassessed at the time of review, regardless of the initial laboratory interpretation. Molecular diagnoses were verified using the current American College of Medical Genetics and Genomics/Association for Molecular Pathology interpretation guidelines for sequence variants^30^ or copy number variants^31^ as appropriate. Considering the established prognostic implications of TSC and variants in genes related to GATOR complex^13,14^, we separated people with these disorders into mTORopathies and other genetic etiologies. The individuals who underwent epilepsy surgery and were not identified as having potentially genetic epilepsy by any of the screening strategies outlined above were included in the non-genetic epilepsy group.

### Clinical variables

Clinical data for both cohorts was derived from the in-house IRB-approved Epilepsy Center Outcomes Registry (IRB ID 17-1015), which captures all patients who undergo a surgical evaluation in Cleveland Clinic Epilepsy Center program. We obtained the following variables: i) demographic (sex, age of seizure onset, preoperative seizure frequency); ii) surgery-related (age at intervention, type of intervention, side of intervention); iii) outcome-related (length of follow-up, time to seizure recurrence (if applicable), and Engel score at last follow-up); iv) clinical (presence of MRI lesion, presence of developmental delay, interictal and ictal electroencephalography (EEG) characteristics, invasive evaluation). We grouped surgery types into temporal, extratemporal, multilobar, and hemispherectomy categories. Individuals with missing data were excluded from the analysis; no data imputation was performed.

We aimed to investigate long-term seizure outcomes and chose 5 years as a time point commonly utilized in epilepsy outcome research as a long-term follow-up milestone.^2,32^ We assigned seizure freedom status five years after surgery based on the time to seizure recurrence. Seizure freedom was defined as Engel score Ia, seizure reduction was any Engel score except Engel score IV, and favorable outcome was defined as Engel score I or II.

### Matching

We additionally performed a matched analysis to level out the possible influence of the clinical epilepsy phenotype on postsurgical outcomes and test the reproducibility of results derived from the multivariate analysis. We matched individuals with non-genetic epilepsy separately to individuals with mTORopathies and other genetic epilepsy etiologies using 1:1 nearest-neighbor matching on propensity scores. Propensity scores were derived by running a generalized linear regression model where the dependent variable was the genetic status, and predictors included clinical variables known to be associated with postsurgical outcomes^33,34^: sex, age at seizure onset and surgery, presence of MRI lesion, presence of developmental delay, preoperative seizure frequency, EEG findings at presurgical evaluation, and side and extent of intervention. Individuals with mTORopathies were matched with individuals with non-genetic epilepsies first. Matched individuals with non-genetic epilepsy were excluded from the second round of matching for individuals with genetic epilepsies other than mTORopathies. Matching was performed with R programming language (ver. 4.2.0) via RStudio graphical user interface (2022.12.0, build 353) using the MatchIt package.

### Statistical analysis

We reported descriptive statistics using ratios, medians, and interquartile ranges (IQR). Considering the non-parametric nature of data, we used the Kruskal-Wallis and Fisher tests to compare continuous and categorical variables, respectively. We applied Bonferroni correction when performing multiple testing at a significance level of α=0.05. We used Kaplan-Meier survival analysis to calculate longitudinal cumulative seizure recurrence risk at five years and Cox proportional hazards models for univariate and multivariate analysis. Statistical analysis and visualization were performed with R programming language (ver. 4.2.0) via RStudio graphical user interface (2022.12.0, build 353).

### Standard Protocol Approvals, Registrations, and Patient Consents

The study was approved by the Cleveland Clinic’s Institutional Review Board as an informed consent-exempt study (protocol identifiers 23-251 and 17-1015).

### Data Availability

The data supporting the findings of this study are available within the article and its supplementary material. Anonymized data not contained in the article or supplemental materials may be shared by request from any qualified investigator.

## Results

### The clinical features of people with genetic epilepsy who received epilepsy surgery differ from those of people with non-genetic epilepsy

We included 585 individuals with non-genetic epilepsy and 49 individuals with genetic epilepsies, including 32 individuals with mTORopathies (5 with GATOR1-related disorders and 28 with tuberous sclerosis) and 17 individuals with epilepsies due to other genetic etiologies who satisfied the study inclusion criteria (Supplemental Figure 1). Out of 28 individuals with genetic epilepsies diagnosed based on genetic test results (11 individuals with mTORopathies and all individuals with other genetic etiologies), for 19 (68%) individuals, the genetic test result was available at the time of surgical decision-making (Supplemental Figure 2).

We observed significant differences in the clinical presentation of individuals in all three groups after correction for multiple comparisons (Table 1 and Figure 1). mTORopathies group had the earliest seizure onset (median 0.3, IQR 0.2-2.0 years) compared to individuals with other genetic (median 4.0, IQR 1.5-12 years) and non-genetic epilepsy etiologies (median 12.0, IQR 4.0-26.0 years). Age of onset was lower in mTORopathies (median 6.0, IQR 3.0-9.5 years) and the group of other genetic etiologies (median 10.0, IQR 5.0-19.0 years) than in individuals with non-genetic etiology (median 32.2, IQR 20.6-46.2 years).

**Figure 1.**
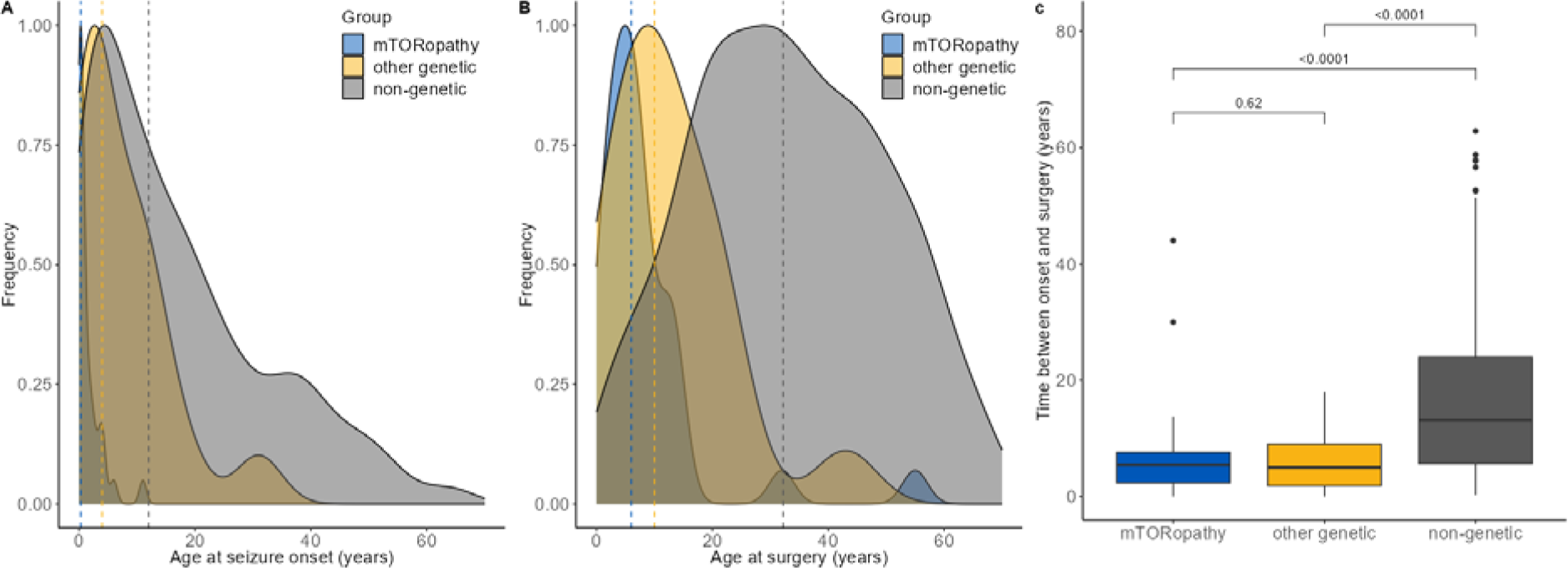
Temporal characteristics of seizure onset and surgical treatment in individuals with germline genetic epilepsies (separated into mTORopathies and other genetic disorders) and non-genetic epilepsies. **A-B**. Density plots of age of seizure onset (A) and at surgery (B) in all three groups. Individuals with epilepsy attributed to mTORopathies and other genetic disorders have a younger age of onset and age at surgery than individuals with non-genetic epilepsy. **C.** Box plot of time between seizure onset and surgery in all three groups. Individuals with mTORopathies and other genetic disorders had a smaller time gap between seizure onset and epilepsy surgery than individuals with non-genetic epilepsy.

**Table 1.**
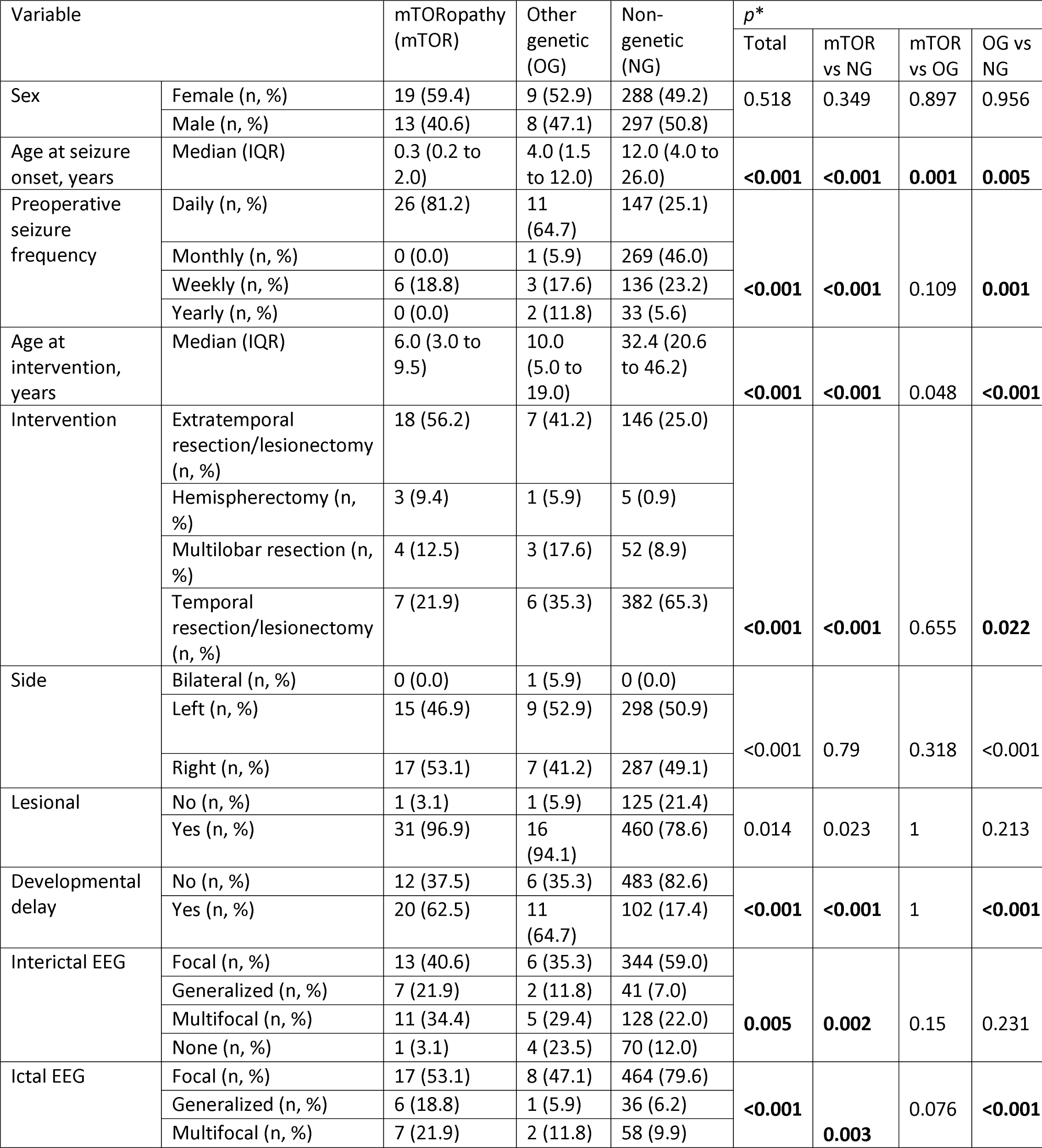
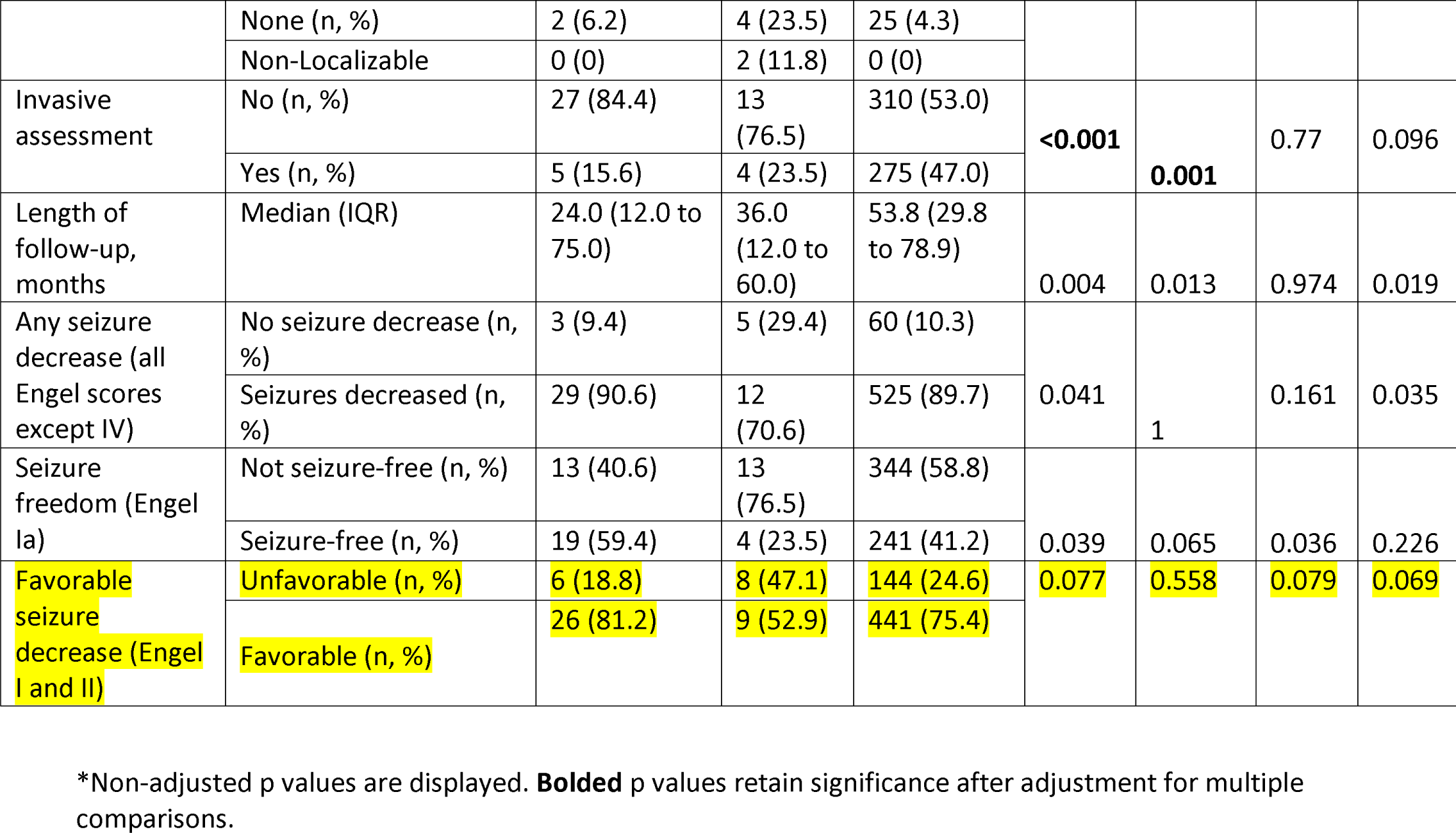
Demographic and clinical cohort comparison.

| Variable |  | mTORopathy (mTOR) | Other genetic (OG) | Non-genetic (NG) | *p* * |  |  |  |
| --- | --- | --- | --- | --- | --- | --- | --- | --- |
|  |  |  |  |  | Total | mTOR vs NG | mTOR vs OG | OG vs NG |
| Sex | Female (n, %) | 19 (59.4) | 9 (52.9) | 288 (49.2) | 0.518 | 0.349 | 0.897 | 0.956 |
|  | Male (n, %) | 13 (40.6) | 8 (47.1) | 297 (50.8) |  |  |  |  |
| Age at seizure onset, years | Median (IQR) | 0.3 (0.2 to 2.0) | 4.0 (1.5 to 12.0) | 12.0 (4.0 to 26.0) | **<0.001** | **<0.001** | **0.001** | **0.005** |
| Preoperative seizure frequency | Daily (n, %) | 26 (81.2) | 11 (64.7) | 147 (25.1) | **<0.001** | **<0.001** | 0.109 | **0.001** |
|  | Monthly (n, %) | 0 (0.0) | 1 (5.9) | 269 (46.0) |  |  |  |  |
|  | Weekly (n, %) | 6 (18.8) | 3 (17.6) | 136 (23.2) |  |  |  |  |
|  | Yearly (n, %) | 0 (0.0) | 2 (11.8) | 33 (5.6) |  |  |  |  |
| Age at intervention, years | Median (IQR) | 6.0 (3.0 to 9.5) | 10.0 (5.0 to 19.0) | 32.4 (20.6 to 46.2) | **<0.001** | **<0.001** | 0.048 | **<0.001** |
| Intervention | Extratemporal resection/lesionectomy (n, %) | 18 (56.2) | 7 (41.2) | 146 (25.0) | **<0.001** | **<0.001** | 0.655 | **0.022** |
|  | Hemispherectomy (n, %) | 3 (9.4) | 1 (5.9) | 5 (0.9) |  |  |  |  |
|  | Multilobar resection (n, %) | 4 (12.5) | 3 (17.6) | 52 (8.9) |  |  |  |  |
|  | Temporal resection/lesionectomy (n, %) | 7 (21.9) | 6 (35.3) | 382 (65.3) |  |  |  |  |
| Side | Bilateral (n, %) | 0 (0.0) | 1 (5.9) | 0 (0.0) | **<0.001** | 0.79 | 0.318 | **<0.001** |
|  | Left (n, %) | 15 (46.9) | 9 (52.9) | 298 (50.9) |  |  |  |  |
|  | Right (n, %) | 17 (53.1) | 7 (41.2) | 287 (49.1) |  |  |  |  |
| Lesional | No (n, %) | 1 (3.1) | 1 (5.9) | 125 (21.4) | 0.014 | 0.023 | 1 | 0.213 |
|  | Yes (n, %) | 31 (96.9) | 16 (94.1) | 460 (78.6) |  |  |  |  |
| Developmental delay | No (n, %) | 12 (37.5) | 6 (35.3) | 483 (82.6) | **<0.001** | **<0.001** | 1 | **<0.001** |
|  | Yes (n, %) | 20 (62.5) | 11 (64.7) | 102 (17.4) |  |  |  |  |
| Interictal EEG | Focal (n, %) | 13 (40.6) | 6 (35.3) | 344 (59.0) | **0.005** | **0.002** | 0.15 | 0.231 |
|  | Generalized (n, %) | 7 (21.9) | 2 (11.8) | 41 (7.0) |  |  |  |  |
|  | Multifocal (n, %) | 11 (34.4) | 5 (29.4) | 128 (22.0) |  |  |  |  |
|  | None (n, %) | 1 (3.1) | 4 (23.5) | 70 (12.0) |  |  |  |  |
| Ictal EEG | Focal (n, %) | 17 (53.1) | 8 (47.1) | 464 (79.6) | **<0.001** | **0.003** | 0.076 | **<0.001** |
|  | Generalized (n, %) | 6 (18.8) | 1 (5.9) | 36 (6.2) |  |  |  |  |
|  | Multifocal (n, %) | 7 (21.9) | 2 (11.8) | 58 (9.9) |  |  |  |  |
|  | None (n, %) | 2 (6.2) | 4 (23.5) | 25 (4.3) |  |  |  |  |
|  | Non-Localizable | 0 (0) | 2 (11.8) | 0 (0) |  |  |  |  |
| Invasive assessment | No (n, %) | 27 (84.4) | 13 (76.5) | 310 (53.0) | **<0.001** | **0.001** | 0.77 | 0.096 |
|  | Yes (n, %) | 5 (15.6) | 4 (23.5) | 275 (47.0) |  |  |  |  |
| Length of follow-up, months | Median (IQR) | 24.0 (12.0 to 75.0) | 36.0 (12.0 to 60.0) | 53.8 (29.8 to 78.9) | 0.004 | 0.013 | 0.974 | 0.019 |
| Any seizure decrease (all Engel scores except IV) | No seizure decrease (n, %) | 3 (9.4) | 5 (29.4) | 60 (10.3) | 0.041 | 1 | 0.161 | 0.035 |
|  | Seizures decreased (n, %) | 29 (90.6) | 12 (70.6) | 525 (89.7) |  |  |  |  |
| Seizure freedom (Engel Ia) | Not seizure-free (n, %) | 13 (40.6) | 13 (76.5) | 344 (58.8) | 0.039 | 0.065 | 0.036 | 0.226 |
|  | Seizure-free (n, %) | 19 (59.4) | 4 (23.5) | 241 (41.2) |  |  |  |  |
| Favorable seizure decrease (Engel I and II) | Unfavorable (n, %) | 6 (18.8) | 8 (47.1) | 144 (24.6) | 0.077 | 0.558 | 0.079 | 0.069 |
|  | Favorable (n, %) | 26 (81.2) | 9 (52.9) | 441 (75.4) |  |  |  |  |

\*Non-adjusted p values are displayed. **Bolded** p values retain significance after adjustment for multiple comparisons.

Both genetic groups showed a higher preoperative seizure frequency. Interictal EEG findings did not differ significantly between the groups. Both mTORopathies and the group of other genetic etiologies had a higher rate of non-focal seizure onset localization than the group with non-genetic etiology, as well as a higher rate of extratemporal and extensive resections, such as hemispherectomies and multilobar resections. Expectedly, the developmental delay was more common in mTORopathies (62.5%) and the group of other genetic etiologies (64.7%) groups compared to the group with non-genetic etiologies (17.7%). The majority of individuals in all three groups had an MRI brain lesion.

### Individuals with genetic epilepsy with etiologies other than mTORopathies tend to have worse outcomes than individuals with mTORopathies and non-genetic disorders

A smaller proportion of individuals with other genetic etiologies achieved seizure freedom at the last follow-up (24%) compared to individuals with mTORopathies (59.0%, *p*_unadj_=0.036, *p*_adj_=0.473) and with non-genetic etiology (41.1%, *p*_unadj_=0.226, *p*_adj_=1). Likewise, the rate of seizure reduction was lowest in the group of other genetic etiologies (70.6%) compared to individuals with mTORopathies (91.0%, *p*_unadj_=0.161, *p*_adj_=0.473) and non-genetic etiologies (89.8%, *p*_unadj_=0.035, *p*_adj_=0.39). The rate of favorable outcomes was 52.9% in of individuals with other genetic etiologies, 81.2% in individuals with mTORopathies (p_unadj_=0.079, p_adj_=0.756), and 75.4% in individuals with non-genetic epilepsy (p_unadj_=0.079, p_adj_=0.756).

The longitudinal analysis demonstrated a higher cumulative probability of seizure recurrence in the group of other genetic etiologies (Figure 2) compared to individuals with mTORopathies and non-genetic epilepsy (HR 3.07, 95% CI 1.4-6.74) at 5 years postoperatively. In multivariate analysis, individuals with etiologies other than mTORopathies had 3.81 times higher seizure recurrence risk (95% CI 1.69-8.62, p=0.001) when adjusting for sex, age at seizure onset, age at surgery, presence of MRI lesion, preoperative seizure frequency, type of surgery, and preoperative EEG (Table 2).

**Figure 2.**
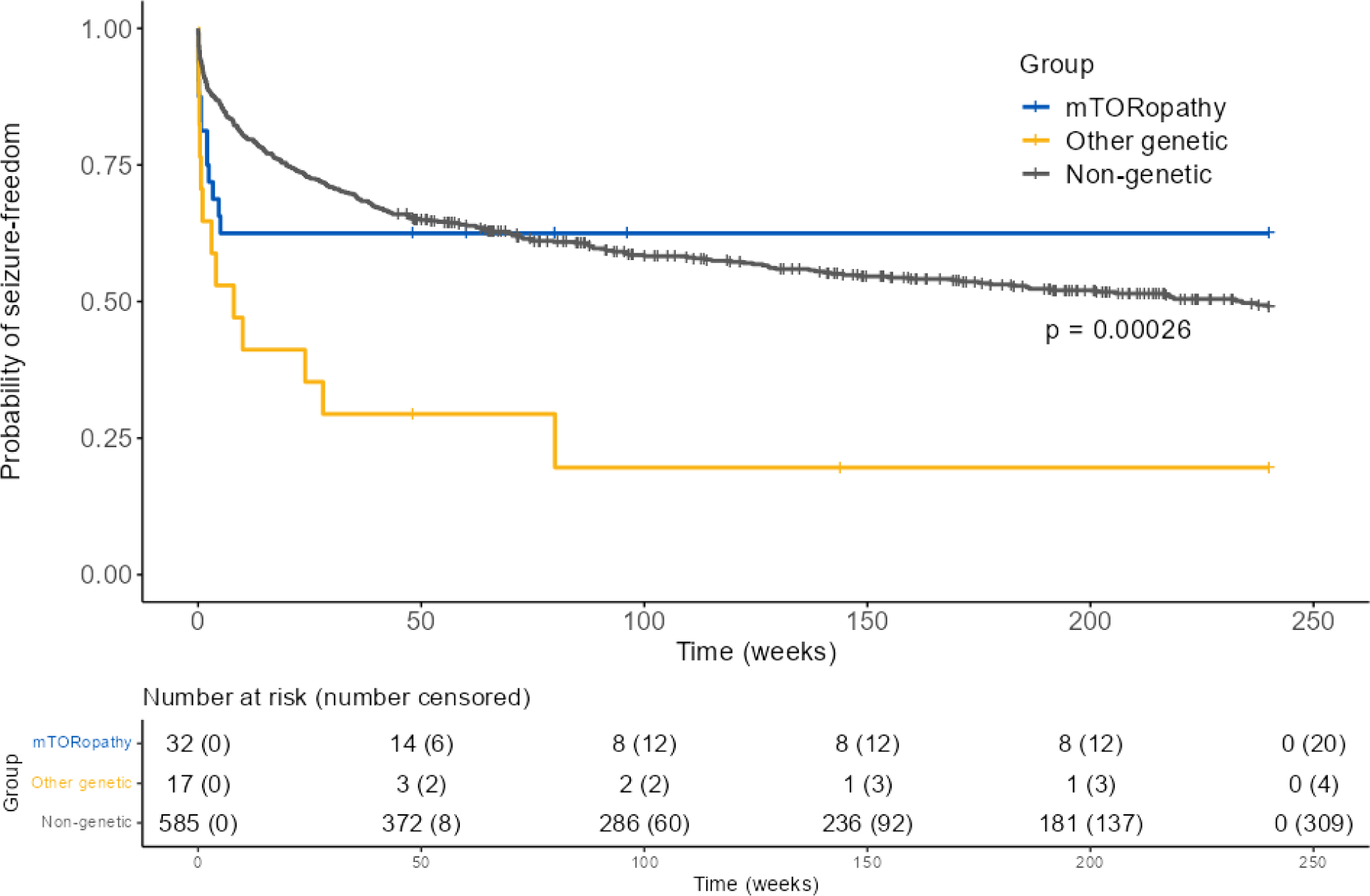
Kaplan-Meier curves of cumulative probability of continuous seizure freedom in unmatched individuals with mTORopathies, other genetic epilepsies, and non-genetic etiologies at 5 years.

**Table 2.** Cox proportional hazards model analysis of association of genetic and non-genetic epilepsy etiology with 5-year risk of seizure recurrence in unmatched and matched genetic and non-genetic epilepsy groups adjusted for clinical variables known to predict postsurgical seizure outcomes (see Methods).

| Variables | Unmatched |  |  | Matched |  |  |
| --- | --- | --- | --- | --- | --- | --- |
|  | HR | 95%CI | p | HR | 95% CI | p |
| mTORopathy | Reference |  |  | Reference |  |  |
| Other genetic | 3.81 | 1.69-8.62 | 0.001 | 2.27 | 1.19-11.60 | 0.02 |
| Non-genetic | 1.11 | 0.59-2.10 | 0.74 | - | - | - |
| Non-genetic – matched mTORopathies | - | - | - | 0.52 | 0.21-1.26 | 0.15 |
| Non-genetic – matched to other genetic | - | - | - | 0.34 | 0.08-1.32 | 0.12 |

We repeated the analysis in a propensity scores-matched sub-cohort (32 individuals with non-genetic epilepsy matched to 32 individuals with mTORopathies and 17 individuals with non-genetic epilepsy matched to 17 individuals with other genetic etiologies; clinical characteristics of the matched sub-cohorts are given in Supplementary Table 3). Similar to non-matched analysis, individuals with genetic etiologies other than mTORopathies had the highest cumulative probability of seizure recurrence at 5 years after surgery among all groups, including the respectively matched group of individuals with non-genetic epilepsy (HR 2.7, 95%CI 1.12-6.67, p=0.02; Figure 3), which persisted in the multivariate analysis (HR 5.8, 95%CI 1.81-19.04, p=0.003; Table 2 and Supplementary Table 3).

**Figure 3.**
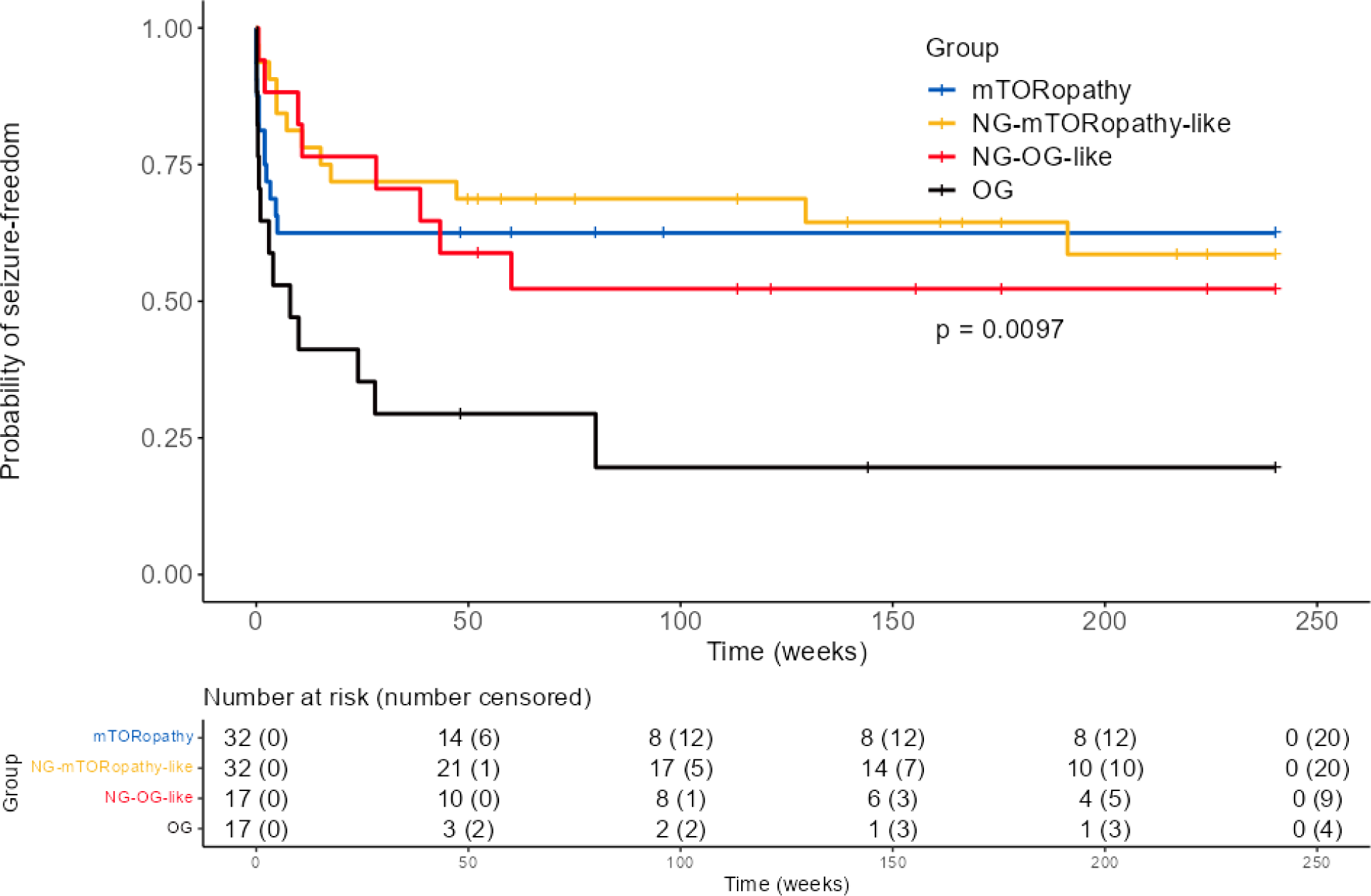
Kaplan-Meier curves of cumulative probability of continuous seizure freedom in matched individuals with mTORopathies, other genetic epilepsies, and non-genetic etiologies at 5 years. Individuals with mTORopathies and epilepsy due to other genetic disorders were each matched to individuals with non-genetic epilepsy 1:1 as specified in the Methods. Individuals with genetic epilepsy other than mTORopathy have the highest risk of seizure recurrence at 5 years, while the risk in respective matched individuals is similar to that in mTORopathies and the corresponding matched group. NG, non-genetic; OG, other genetic.

## Discussion

With the expanding use of genetic testing in the diagnostic workup of individuals with epilepsy and the increasing spectrum of genetic epilepsy phenotypes described, determining the surgical eligibility of individuals with genetic epilepsy has become an imperious clinical need. Our study investigated epilepsy-related phenotypes and outcomes of 49 surgically treated individuals with genetic epilepsies, including 32 individuals with mTORopathies and 17 individuals with other genetic etiologies, compared to those of 585 individuals with non-genetic epilepsy. We identified a range of differences in epilepsy surgery-oriented phenotypes in all three groups. We also showed that individuals with genetic epilepsies, due to the variants in genes other than mTOR pathway genes, tend to have worse outcomes in terms of seizure freedom but can still attain seizure reduction.

The presence of developmental delay was a differentiating feature of individuals with genetic epilepsies in our cohort. Indeed, developmental delay of different severity is a feature of numerous genetic disorders associated with epilepsy. The yield of genetic testing in individuals with epilepsy increases if concurrent developmental delay is present^33,34^, making this combination of clinical features an important triaging factor for genetic testing.^35^ We thus suggest that the presence of developmental delay in individuals assessed for epilepsy surgery prompts consideration of genetic testing.

Most of the individuals from both genetic and non-genetic groups in our study had epilepsy attributed to an MRI lesion. The bias towards lesional cases is expected in the overall patient population seen at an epilepsy surgery program and is attributed to the known role of the presence of a lesion in postsurgical seizure outcomes. ^36^ However, this is especially interesting with respect to surgical decision-making in conjunction with the timing of genetic testing in individuals with genetic epilepsies. In the majority of individuals with genetic epilepsy in our cohort, results of genetic testing were available at the time of surgical intervention. This can suggest that conventional surgical candidacy determinants rather than genetic status were the major drivers for surgical decision-making.

In our study, individuals with mTORopathies, which included tuberous sclerosis complex (TSC) and GATOR-related (caused by variants in *DEPDC5*, *NPRL3*, and *NPRL2*) disorders, had favorable outcomes, with 59.4% of individuals being seizure-free at the time of last follow-up. Large multicenter studies have established a good short- and long-term prognosis in individuals with TSC with seizure freedom ranging from 50% to 70% depending on the assessment points and clinical factors, such as including the dominant tuber in the resection or more extensive resections.^11–13^ Favorable outcomes in GATOR1-related disorders are in line with an available evidence. GATOR1-related epilepsy disorders have only been delineated recently,^37,38^ and the data on the seizure outcomes in these disorders is limited to case series, which show an overall favorable seizure-related prognosis ^14,24,27,28^ with 92.6% of seizure freedom reported in the most extensive to-date single-center study.^15^ Interestingly, in our study, although not statistically significant, the rate of seizure freedom in mTORopathies was higher than in people with non-genetic epilepsy, and the cumulative risks of seizure recurrence at 5 years were comparable to those of individuals with non-genetic etiology. Epilepsy in both tuberous sclerosis and GATOR1-related disorders is caused by malformations of cortical development, which include cortical and subcortical tubers and focal cortical dysplasias in tuberous sclerosis and a range of malformations ranging from MRI-negative focal cortical dysplasias to hemimegalencephaly in GATOR1-related epilepsies.^3,7^ In both cases, the focality of the malformations of cortical development is attributed to somatic second hit variants restricted to the lesions.^9,10^ Focality of epilepsy in mTORopathies caused by a common biological mechanism re-enforces amenability of these disorders to surgery, which can be expected to be on a par with focal non-genetic epilepsies.

In contrast, individuals with other generic etiologies showed less favorable surgical prognosis with a lower rate of seizure freedom at the last follow-up and higher hazards of seizure recurrence at 5 years. The available literature on epilepsy surgery outcomes in non-mTOR-related genetic epilepsy disorders is likewise restricted and reports heterogeneous outcomes depending on the exact type of genetic disorder. ^8,13,14,24,27,28^ So far, ion channel and synaptic transmission disorders have been associated with poor postsurgical outcomes even in the presence of an MRI-lesion^23,24^. In contrast, disorders related to copy number variants (CNVs) appear to respond to surgical treatment^24,39^. We did not identify any individuals with ion channel-related disorders who underwent resective epilepsy surgery in our center (Supplemental Table 4). One patient with *PCDH19*-related disorder, a disorder related to synaptic transmission, had an unfavorable outcome at the last follow-up. Likewise, out of 6 individuals with CNVs, 4 did not achieve seizure freedom, and 2 had seizure worsening postoperatively (Engel score IV). It is worth noting that more than 70% of individuals in the group of other genetic etiologies still achieved seizure reduction. Although seizure freedom is the ultimate goal of epilepsy surgery^40^ and nearly universal patients’ and caregivers’ expectation^41^, it is often not achievable regardless of the etiology for various reasons: multifocality, failure to identify seizure onset zone, localization of the focus in the eloquent area and other factors^42,43^. Furthermore, genetic epilepsies are frequently associated with a high seizure burden that is not controlled by medications and significantly impacts both patients and their caregivers’ quality of life^44,45^. Hence, before etiology-targeted precision therapies enter the medical market, palliative seizure reduction with resective surgery could be a reasonable goal of care in specific clinical scenarios for individuals with genetic epilepsies. More data on surgical outcomes in specific genetic etiologies, as well as their comparison to other invasive approaches such as neurostimulation, is needed to optimize the use of available methods for treating patients with genetic epilepsies.

Numerous studies have explored predictors of seizure-related outcomes in different types of surgeries using non-invasive and invasive electroclinical data^2,32,46,47^. While there is a continuing effort to account for transcriptomics, single nucleotide polymorphism, and somatic variant data with regard to postsurgical outcome predictions^48–50^, clinically accessible germline genetic etiology has received little attention. In our multivariable analysis adjusting for the known predictors of outcome, genetic etiology was a significant factor affecting the cumulative 5-year risk of seizure recurrence. Although the grouping of genetic etiologies is crude, this preliminary evidence suggests that genetic etiology can serve as a biomarker of seizure outcomes and be embedded in future seizure outcome prediction algorithms.

The main limitation of our study is the small size of the genetic epilepsy cohort. Yet, this is one of the largest to-date reported single-center genetic cohorts of surgically treated individuals with germline genetic epilepsy. The single-center setting impacts the generalizability of our findings but also decreases bias from interinstitutional differences in epilepsy surgery practice and allows for a structured follow-up and unified outcome assessment. We acknowledge that some non-genetic cases may have an undiagnosed genetic etiology. To mitigate this bias, we removed individuals identified as harboring potentially genetic epilepsy identified through our EMR search.

In summary, the clinical differences we found in the gross comparison of the cohorts delineated the phenotype of a potential surgical candidate with genetic epilepsy. We quantified the seizure-related outcomes of epilepsy surgery in individuals with germline genetic epilepsy deemed to be surgical candidates in comparison to individuals with non-genetic epilepsy. Within constraints of the study limitations, our results suggest that the seizure-related outcomes in epilepsies due to genetic etiologies other than mTORopathies tend to be worse than in mTORopathies and non-genetic epilepsy cases. Still, the resulting seizure decrease can be reasonable depending on the goals of care. Our results support the role of genetic etiology as a biomarker of postsurgical seizure-related outcomes. Further studies are needed to validate these findings and clarify the value of different genetic etiologies in predicting postsurgical epilepsy prognosis.

## Data Availability

All data produced in the present study are available upon reasonable request to the authors.

