## Supplementary Figures for "Evaluating Clinical Presentation and Long-Term Outcomes in Individuals with Genetic and Non-Genetic Epilepsy Treated with Epilepsy Surgery: A Single-Center Study"

### Supplementary Figure 1: Patient selection flowchart

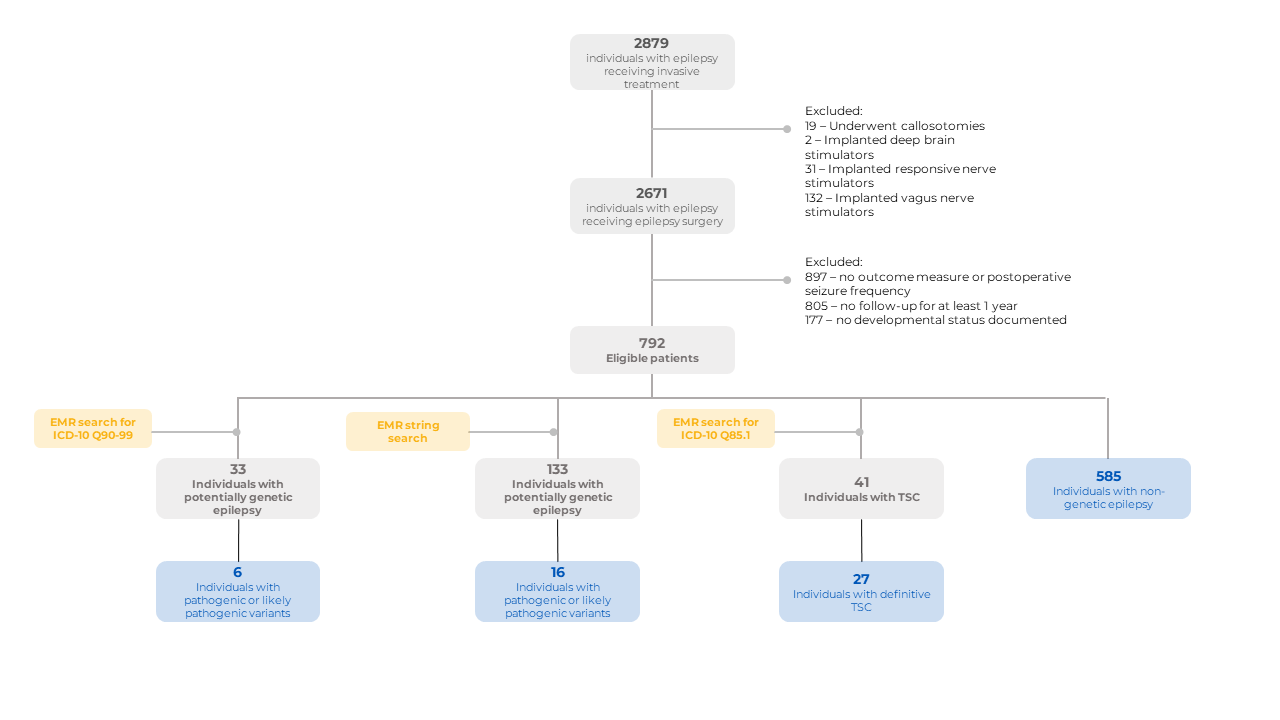

### Supplementary Figure 2: Genetic testing in individuals with genetic epilepsy who underwent epilepsy surgery at the Cleveland Clinic Epilepsy Center

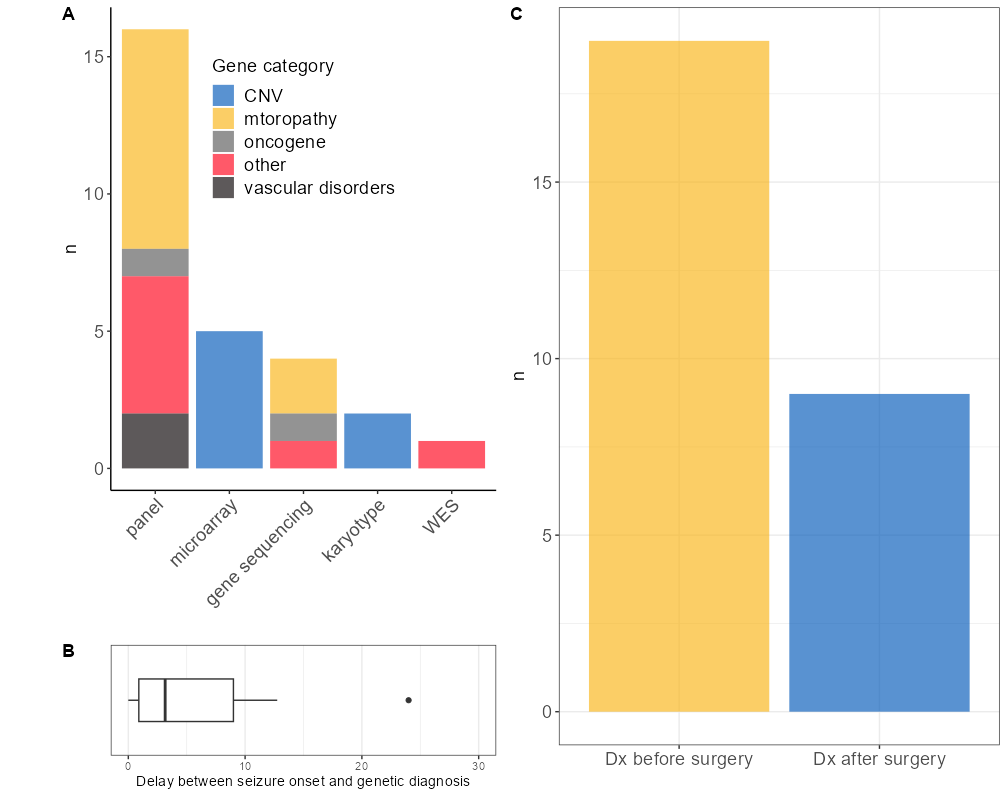

Figure S2. A. Barplot of types of genetic testing used for genetic diagnosis in cohort people with genetic epilepsy. Most individuals were diagnosed with genetic panels. B. Box plot of time gap between seizure onset and genetic diagnosis. For 3 individuals, genetic diagnosis was known before the seizure onset (individuals with 16p13.11 microdeletion, partial trisomy 18, and *NAGLU*-related disorder). C. Barplot of genetic status availability before surgery. In 19/28 (68%) individuals, genetic diagnosis was available before surgery.
